# Improved Seizure Onset-Zone Lateralization in Temporal Lobe Epilepsy using 7T Resting-State fMRI: A Direct Comparison with 3T

**DOI:** 10.1101/2023.06.06.23291025

**Authors:** Alfredo Lucas, Eli J. Cornblath, Nishant Sinha, Lorenzo Caciagli, Peter Hadar, Ashley Tranquille, Joel M. Stein, Sandhitsu Das, Kathryn A. Davis

**Affiliations:** Perelman School of Medicine, University of Pennsylvania; Department of Bioengineering, University of Pennsylvania; Department of Neurology, University of Pennsylvania; Department of Neurology, Massachussets General Hospital (work conducted while at the University of Pennsylvania); Department of Radiology, University of Pennsylvania

## Abstract

**Objective:** Resting-state functional magnetic resonance imaging (rs-fMRI) at ultra high-field strengths (≥7T) is known to provide superior signal-to-noise and statistical power than comparable acquisitions at lower field strengths. In this study, we aim to provide a direct comparison of the seizure onset-zone (SOZ) lateralizing ability of 7T rs-fMRI and 3T rs-fMRI.

**Methods:** We investigated a cohort of 70 temporal lobe epilepsy (TLE) patients. A paired cohort of 19 patients had 3T and 7T rs-fMRI acquisitions for direct comparison between the two field strengths. Forty-three patients had only 3T, and 8 patients had only 7T rs-fMRI acquisitions. We quantified the functional connectivity between the hippocampus and other nodes within the default mode network (DMN) using seed-to-voxel connectivity, and measured how hippocampo-DMN connectivity could inform SOZ lateralization at 7T and 3T field strengths.

**Results:** Differences between hippocampo-DMN connectivity ipsilateral and contralateral to the SOZ were significantly higher at 7T (p_FDR_=0.008) than at 3T (p_FDR_=0.80) when measured in the same subjects. We found that our ability to lateralize the SOZ, by distinguishing subjects with left TLE from subjects with right TLE, was superior at 7T (AUC = 0.97) than 3T (AUC = 0.68). Our findings were reproduced in extended cohorts of subjects scanned at either 3T or 7T. Our rs-fMRI findings at 7T, but not 3T, are consistent and highly correlated (Spearman Rho=0.65) with clinical FDG-PET lateralizing hypometabolism.

**Significance:** We show superior SOZ lateralization in TLE patients when using 7T relative to 3T rs-fMRI, supporting the adoption of high-field strength functional imaging in the epilepsy presurgical evaluation.

## Introduction

Functional magnetic resonance imaging (fMRI) is the main non-invasive approach for assessing whole brain function *in vivo*. In epilepsy, task-based fMRI at field strengths of 1.5-Tesla (T) and 3T are used clinically for language lateralization prior to surgical resection^1^. Resting-state fMRI (rs-fMRI), however, remains yet to be clinically translated to the presurgical evaluation of epilepsy, despite studies at 3T demonstrating potential for seizure onset zone (SOZ) lateralization^2–4^. Small sample sizes and low statistical power remain limiting factors in many of these studies^5^, preventing the detection of meaningful differences at a single subject level required for eventual translation to the clinic.

In healthy subjects, task-based fMRI studies at ultra-high magnetic field strengths (≥7T) have shown superior temporal signal-to-noise ratio (tSNR), and superior functional contrast to noise ratio (fCNR), when compared to equivalent 3T acquisitions^6–10^. Further, some of these studies have shown that 7T acquisitions also lead to at least a triplication of statistical power compared to an equivalent 3T acquisition^8, 9^. In clinical cohorts, however, few functional neuroimaging studies have directly compared 3T and 7T acquisitions^11, 12^.

Many rs-fMRI studies in epilepsy at 3T have found functional changes that suggest network reorganization^2–4, 13, 14^. For example, decreased ipsilateral hippocampal connectivity to the default mode network (DMN) remains the most commonly reproduced finding in case-control studies of temporal lobe epilepsy (TLE)^15–20^. At 7T, rs-fMRI studies have also uncovered functional network reorganization in epilepsy^21–23^, but none have directly compared the utility of 3T vs. 7T fMRI in describing the underlying epileptogenic networks, or lateralizing the SOZ. This is in contrast to the many studies comparing the clinical utility of 7T and 3T structural neuroimaging in epilepsy^24–29^.

In this study, we aimed to address the gap in knowledge regarding the clinical utility of 7T rs-fMRI when compared to 3T rs-fMRI in TLE. To do so, we asked whether 7T rs-fMRI could provide better lateralization of the SOZ in focal TLE than 3T rs-fMRI. To answer this question, we investigated (i) a cohort of TLE patients with paired 7T and 3T rs-fMRI acquisitions, allowing us to directly compare findings across field strengths, and (ii) an extended cohort of patients with only 3T or 7T rs-fMRI, allowing for further validation of the findings from the paired cohort. For each patient, and at both field strengths, we measured the hippocampal functional connectivity to other nodes within the DMN. We then compared how the hippocampal-DMN connectivity ipsilateral to the SOZ differed from the connectivity contralateral to the SOZ, a common approach to study SOZ laterality in the TLE literature^15–20^. Finally, we compared the utility of 3T and 7T rs-fMRI in lateralizing the SOZ at a single subject level, and compared the performance of each field strength with the lateralization of the SOZ by quantitative FDG-PET. Our findings provide evidence for superior SOZ lateralizing ability by 7T rs-fMRI, supporting the adoption of high-field strength functional neuroimaging imaging in the epilepsy presurgical evaluation.

## Methods

### Subject Inclusion

Data acquisition for this study was approved by the institutional review board of the University of Pennsylvania. We compiled a dataset consisting of 70 drug-resistant unilateral TLE patients who underwent neuroimaging during presurgical evaluation including resting state functional magnetic resonance imaging (fMRI). Nineteen patients received both 3T and 7T neuroimaging. Thirty-nine only received 3T and 8 only received 7T neuroimaging.

The Penn Epilepsy Surgical Conference (PESC) determined the location of seizure focus for all patients using a multidisciplinary approach, including clinical, neuroimaging, and neurophysiological factors such as seizure semiology, MRI, PET, MEG, scalp EEG, and intracranial EEG. The PESC team consisted of neurologists, neurosurgeons, neuropsychologists, neuroradiologists, and nuclear medicine specialists. Among the 19 patients in the paired 3T-7T cohort, 12 had left-sided lateralization (L-TLE) and 7 had right-sided lateralization (R-TLE). In the 3T replication cohort (19 paired patients plus 43 patients with 3T imaging only), 37 had left-sided lateralization and 25 had right-sided lateralization. In the 7T replication cohort (19 paired patients plus 8 patients with 7T imaging only), 17 had left-sided lateralization and 10 had right-sided lateralization. The study also included 16 age-matched control participants, 8 of whom were investigated with 3T imaging and 8 with 7T imaging. Demographic information, MRI lesional status, final lateralization, and Engel surgical seizure outcomes^30^, where available, are recorded in Table 1.

**Table 1:** Demographic and clinical information for paired 3T-7T cohort as well as extended 3T and 7T cohorts. **Abbreviations: MTS** – Mesial temporal sclerosis. **Engel** – Engel surgical outcome.

Subject Demographics
| Characteristic | Group |  |  |
| --- | --- | --- | --- |
|  | Paired 3T-7T | Extended 3T | Extended 7T |
| **Number of Subjects** | 19 | 62 | 27 |
| **Age** | 35±11 | 38±13 | 36±11 |
| **Female** | 12 | 34 | 15 |
| **MRI Lesional** | 12 | 40 | 17 |
| *MTS* | 8 | 22 | 11 |
| **Lateralization** |  |  |  |
| *Left* | 12/19 | 37/62 | 17/27 |
| *Right* | 7/19 | 25/62 | 10/27 |
| **Surgical Intervention** | 9 | 25 | 11 |
| *Engel I* | 6/9 | 16/25 | 8/11 |
| *Engel II-IV* | 3/9 | 9/25 | 3/11 |
| **Imaging Protocol Version** |  |  |  |
| *A (3T)* | 5/19 | 13/62 | - |
| *B (3T)* | 14/19 | 49/62 | - |
| *C (7T)* | 4/19 | - | 10/27 |
| *D (7T)* | 15/19 | - | 17/27 |

## Image Acquisition

### 3T Image Acquisition

Two different fMRI acquisition protocols were used at 3T (fMRI protocols A and B; see harmonization details in the **Hippocampal Seed-to-Voxel Connectivity** section). The number of subjects that were scanned with each protocol is shown in **Table 1**. fMRI images for protocol A were acquired with 3.0mm isotropic voxel size, echo time (TE)/ repetition time (TR)= 30/500ms, a multiband factor of 6 and a 7-min acquisition time. fMRI images for protocol B were acquired with a 2.0mm isotropic voxel size, TE/TR= 37/800ms, a multiband factor of 6 and a 9-min acquisition time. For both protocols, T1 weighted images were acquired with a sagittal, 208-slice MPRAGE sequence, TE/TR= 2.24/2400ms, inversion time (TI)=1060ms, field-of-view (FOV)= 256mm, with a 0.8 isotropic voxel size. All 3T control subjects were scanned with protocol B.

### 7T Image Acquisition

Two different fMRI acquisition protocols were used at 7T (fMRI protocols C and D; see harmonization details in the **Hippocampal Seed-to-Voxel Connectivity** section). The number of participants that were scanned with each protocol are shown in **Table 1**. fMRI images for protocol C were acquired with 2.0mm isotropic voxel size, echo time TE/ TR= 23.6/1000ms, a multiband factor of 6 and a 7-min acquisition time. fMRI images for protocol D were acquired with a 1.5mm isotropic voxel size, TE/TR= 20/1000ms, a multiband factor of 6 and a 9-min acquisition time. For protocol C, T1-weighted images were acquired with a 176 slice MPRAGE sequence, TE/TR=4.4/2800ms, TI=1500ms, with a 0.8mm isotropic voxel size. For protocol D, T1-weighted images were acquired with a 160 slice, MP2RAGE sequence, TE/TR=1.86/5000ms, TI1/TI2=700/2700ms, with a 1mm isotropic voxel size. Prior to further anatomic and functional preprocessing, we multiplied the INV2 by the UNI images in our MP2RAGE acquisition to get a better brain extraction. All 7T control participants were scanned with protocol C.

### Functional Imaging Processing

We used fMRIPrep^31^ to perform brain extraction and segmentation of individual T1-weighted (T1w) images, registration of task fMRI BOLD volumes to individual T1w and MNI template space, and time-series confound estimation. We used the fMRIPrep output data as our input to the xcpEngine post-processing pipeline for confound regression, demeaning, detrending and temporal filtering^32^. See Lucas et. al. for specific details on the pre-processing pipeline used in this study^33^.

### tSNR Analysis of Controls

To determine if differences in temporal signal to noise ratio (tSNR) could be influencing our results at 3T and 7T, we measured the tSNR in the fMRI acquisitions of the control subjects. We chose to only inspect the tSNR in controls for two main reasons. First, the tSNR is dependent on the variability (standard deviation) of the timeseries at each voxel. Some studies have argued that disease specific processes, as well as cognitive processes, can influence this variability^34^. Furthermore, studies in people with epilepsy have shown that the BOLD signal variance is significantly greater than the signal variance in controls^35^. Second, there are slight differences in acquisition parameters between the 3T and 7T acquisitions of the epilepsy patients which complicate the interpretation of the tSNR comparison^36–38^. On the other hand, the 3T and 7T acquisitions for the control subjects were similar in terms of voxel size.

We measured the tSNR on the minimally preprocessed fMRI images, consisting of the fMRI acquisition transformed into MNI space. To calculate the tSNR at each voxel, we computed the mean of the timeseries and divided it by the standard deviation of the timeseries. For each subject, we measured the average tSNR in the following regions of interest (ROIs): cortex and subcortex, individual subcortical structures, and the posterior cingulate cortex (PCC) and the medial pre-frontal cortex (MPFC). We chose the PCC and MPFC as we define thse regions as nodes within the DMN in our later analysis. All regions were based on the Harvard-Oxford cortical and subcortical atlases^39, 40^, with the PCC and MPFC defined as specified in: *Comparing Left and Right Seed-to-Voxel Maps – Group Level*.

### Hippocampal Seed-to-Voxel Connectivity

The methods described in the following sections were applied to the fully processed 3T and 7T fMRI acquisitions of each subject as well as to the fully processed fMRI acquisitions of the 3T and 7T extended cohorts. A general overview of the methods is shown in Figure 1.

**Figure 1.**
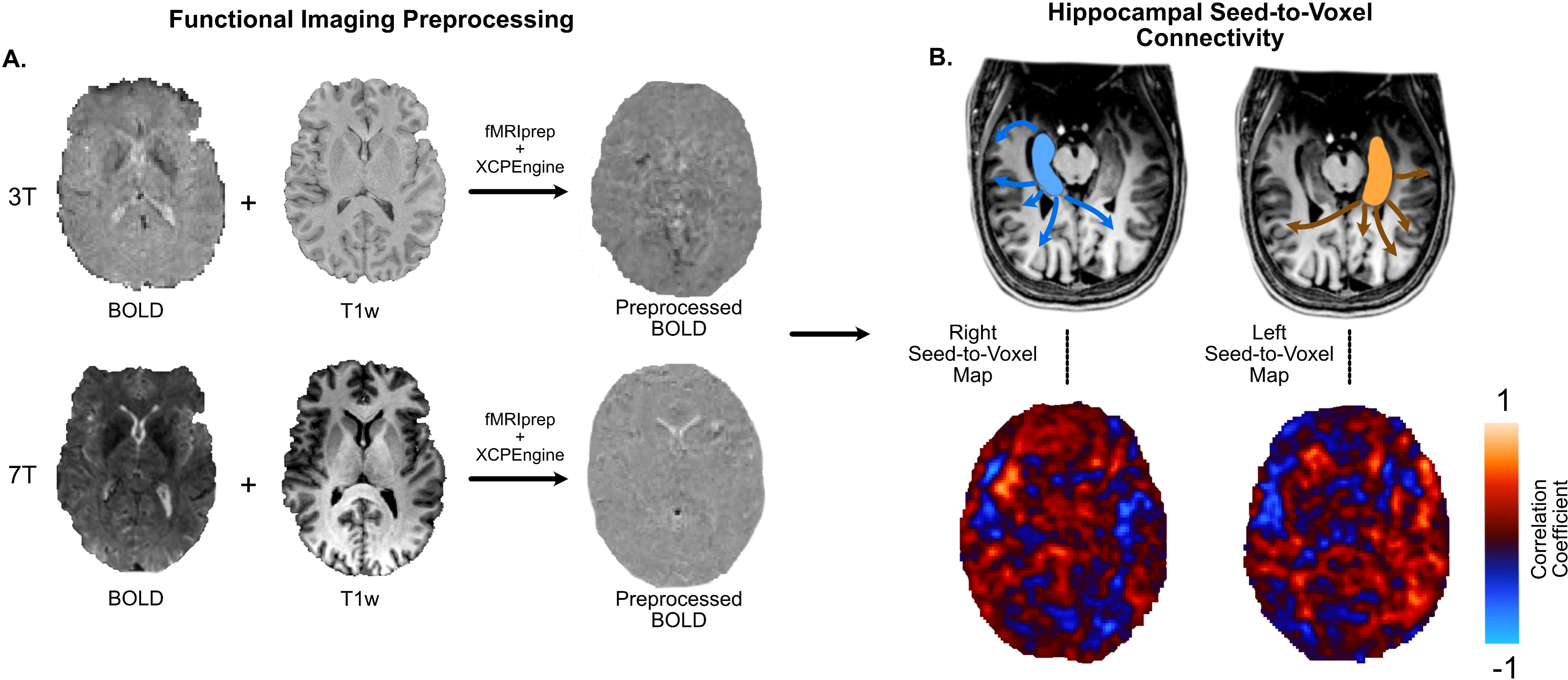
– Methodological Overview. **Panel A** shows the unprocessed BOLD, T1-weighted and processed BOLD images for the same participant at 3T and 7T. Functional imaging processing was carried out for both 3T and 7T with fMRIprep and XCPEngine. For the processed 3T and 7T BOLD images, the hippocampal seed-to-voxel connectivity was computed for the left and right hippocampi as shown in **Panel B**. The left and right seed-to-voxel maps were then used in subsequent analyses throughout the manuscript.

To measure the connectivity of the hippocampus to the DMN, we used seed-to-voxel connectivity^11, 41–43^ (Figure 1B). For each patient, we defined a mask in MNI space for the left and right hippocampi based on the Harvard-Oxford subcortical atlas. We computed the average processed BOLD timeseries within the left and right hippocampal regions of interest (ROIs), which we defined as the left and right seeds respectively. Then, we computed the Pearson’s correlation of the timeseries of each seed ROI, with the timeseries of every other voxel in the brain. After processing, we had two seed-to-voxel maps: one for the left hippocampus as a seed, and one for the right hippocampus as a seed.

Due to differences in the fMRI acquisition protocol potentially influencing the results, we harmonized subjects from different protocols using NeuroCombat^44^. We combined the left and right seed-to-voxel correlation maps for each subject into a matrix, and used it as input for NeuroCombat. The matrix included a batch indicator variable for each subject’s protocol assignment and a group assignment variable indicating whether they were in the left or right TLE group. NeuroCombat removed the batch variable as a covariate while preserving the group assignment variable. We applied this process separately for 3T and 7T acquisitions.

### Comparing Left and Right Seed-to-Voxel Maps – Group Level

To measure whether the difference between left and right seed-to-voxel maps differed between L-TLE and R-TLE, and to assess whether the measured difference was larger at 7T than at 3T, we used voxelwise Cohen’s *d* maps. For each group (L-TLE and R-TLE), we calculated a Cohen’s d map by comparing all right seed-to-voxel maps with all left seed-to-voxel maps. These maps indicate the effect size for each voxel and whether the left or right hippocampal connectivity is larger. Negative Cohen’s d values indicate that the right hippocampus is less connected to a particular voxel than the left hippocampus, while positive values indicate the opposite. We smoothed Cohen’s *d* maps with a Gaussian filter with σ=0.5 voxels for distribution analyses, and with a filter of σ=2 voxels for visualization and correlation analyses.

We measured the distribution of Cohen’s *d* values in two nodes of the default mode network: the posterior cingulate cortex (PCC) and the medial pre-frontal cortex (MPFC) (Figure 2A). These nodes were chosen due to their prominence within the DMN^45^, as well as their previously reported association with TLE^19, 20^. We created PCC and MPFC masks by combining Harvard-Oxford regions and selecting DMN ROI voxels using the Schaefer 200 atlas^46^. The PCC mask included the cingulate gyrus (posterior division) and precuneus cortex, while the MPFC mask included the frontal pole and superior frontal gyrus. We generated separate masks for the left and right hemispheres and measured their distributions in the combined mask.

**Figure 2.**
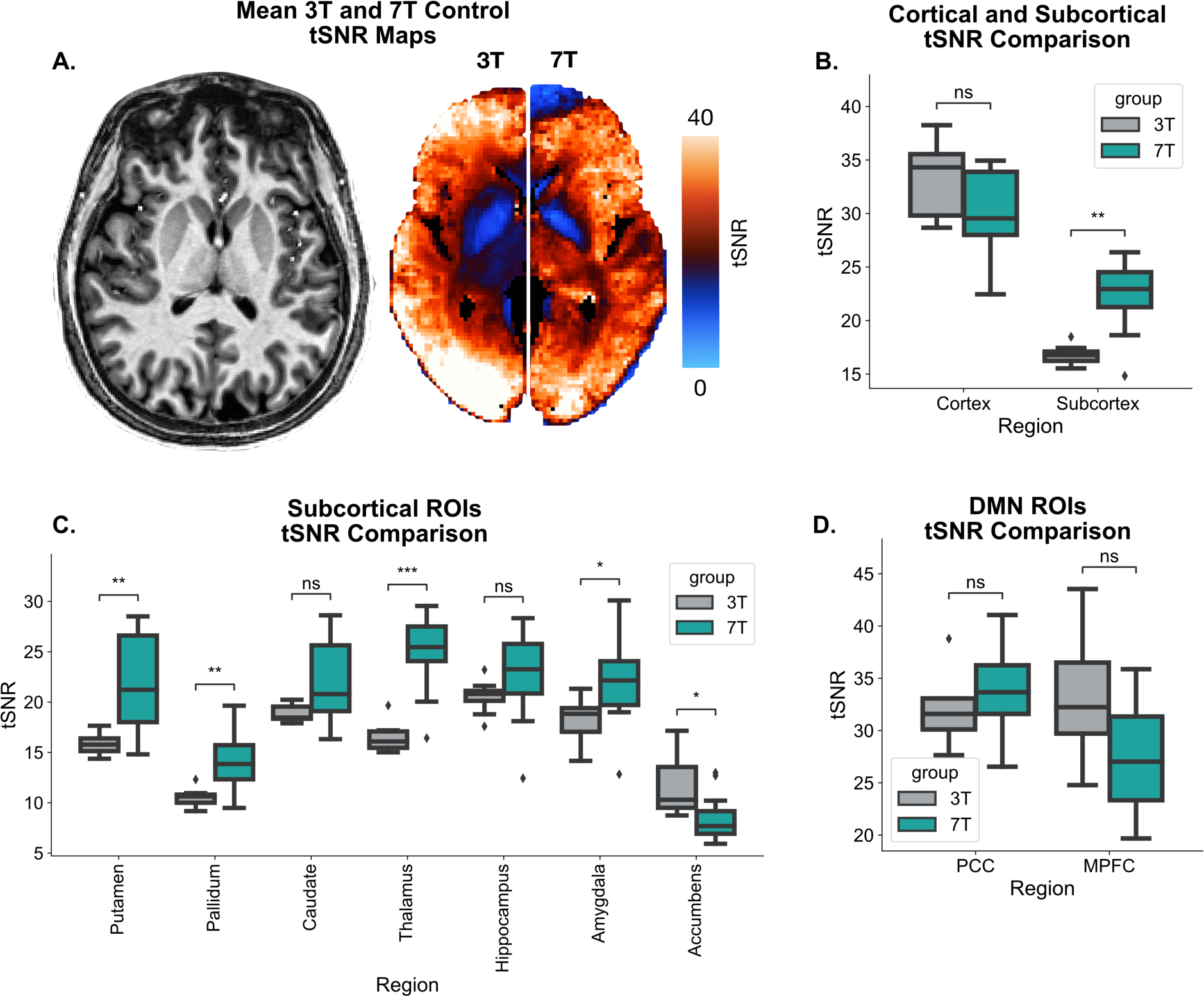
– 3T vs. 7T tSNR Comparison in Controls. In **Panel A**, the left image shows an MP2RAGE T1w acquisition for a representative subject. The right image shows the mean tSNR map for the 3T control subjects and the 7T control subjects side-by-side. **Panel B** shows the mean cortical and subcortical tSNR values, **Panel C** shows the mean tSNR values across all subcortical ROIs, and **Panel D** the mean tSNR values for the PCC and MPFC in the DMN, for 3T and 7T control subjects. *: 0.01 < p <= 0.05, **: 0.001< p <=0.01, ***: 0.0001 < p <= 0.001.

### Comparing Left and Right Seed-to-Voxel Maps – Subject Level

To measure the ability of 3T and 7T rs-fMRI to lateralize the SOZ at a single subject level, we computed for each subject the mean value of the difference between smoothed (Gaussian filter with σ=0.5 voxels) right and left seed-to-voxel hippocampal maps in the posterior cingulate cortex (PCC). For this analysis, we restricted the measurement to the PCC because it was the region where we observed the strongest effect in the group level analysis.

### Quantitative FDG-PET Analysis

Since FDG-PET remains the neuroimaging clinical standard for SOZ lateralization in TLE^47^, we compared quantitative FDG-PET hypometabolism to our rs-fMRI findings at 3T and 7T. Eighteen of the 19 paired subjects had clinical PET scans available for this analysis. For each subject, FDG-PET scans were registered through a rigid transformation to the T1w MRI. Then, we generated subject specific parcellations of the Desikan-Killiany-Tourville (DKT) atlas using FreeSurfer on the T1w image^48^. From these parcellations, we measured the asymmetry index (right-left/right+left) of the FDG-PET contrast within a temporal (DKT labels corresponding to the superior, middle and inferior temporal giri) and a mesial temporal ROI (DKT labels corresponding to the hippocampus, amygdala, parahippocampus and entorhinal cortex). The caudal aspect of the middle frontal gyrus (near the supplementary motor area) was used as a control region.

### Statistical Analysis

We compared tSNR differences between control 3T and 7T acquisitions using a non-parametric Mann-Whitney-U test.

To compare the distribution of Cohen’s *d* values in the PCC and MPFC between L- and R-TLE, we performed a permutation test of the difference between the mean of the L-TLE and R-TLE Cohen’s *d* distributions. We carried out 1000 permutations of the difference between the mean Cohen’s *d* in L-TLE and R-TLE, and the *p*-value for the difference was determined as the number of mean differences in the permuted distribution above the actual mean difference, divided by the total number of permutations, with a positive bias of one in both the numerator and the denominator^49^. Additional comparisons between L-TLE and R-TLE were done using two-sample, two-tailed t-tests. For all analyses, statistical significance was determined at p < 0.05, and Bonferroni correction was used to correct for multiple comparisons where specified (*p_FDR_)*. Reported Cohen’s *d* values are absolute value, and we classify Cohen’s *d* values smaller than 0.5 as small effect sizes. For confidence interval estimates of the area under of receiver operating characteristics curve (ROC AUC) curves the DeLong approximation was used^50^.

## Results

### tSNR of 7T is higher in the subcortical structures

We measured temporal signal-to-noise ratio (tSNR) maps from the rs-fMRI acquisitions of controls acquired at 3T and 7T. The group average tSNR map for healthy controls at 3T and 7T (Figure 2A) shows a similar distribution of voxelwise tSNR values across the brain (Pearson’s r = 0.94, p<0.001). Overall, the tSNR in subcortical ROIs was significantly higher for 7T than for 3T (*p_FDR_*<0.001) (Figure 2B), while there were no significant differences between 3T and 7T in terms of average cortical tSNR, although the value was slightly larger for 3T (*p_FDR_*=0.075). Within specific subcortical ROIs (Figure 2C), the tSNR for 7T was higher in the putamen (*p_FDR_*=0.002), pallidum (*p_FDR_*=0.009), thalamus (*p_FDR_*<0.001) and amygdala (*p_FDR_*=0.015). The nucleus accumbens was the only region to exhibit higher tSNR at 3T (*p_FDR_*=0.015). Within cortical DMN ROIs, no statistically significant differences were seen between 3T and 7T. Overall, our results suggest increased subcortical tSNR at 7T, but a comparable whole cortex tSNR between 3T and 7T.

### Lateralizing group-level effects are stronger at 7T than 3T

To assess effect size differences between 3T and 7T rs-fMRI in quantifying hippocampus-DMN connectivity in TLE, we measured group-level Cohen’s *d* maps between left and right seed-to-voxel maps (Figure 3). At 7T, the inspected DMN regions showed opposite effects in L-TLE and R-TLE. For L-TLE patients, the voxelwise Cohen’s *d* was positive within the PCC and MPFC suggesting that the functional connectivity of these regions to the left hippocampus was lower than their connectivity to the right hippocampus (Figure 3C, Figure 4A). For R-TLE patients, the opposite effect was seen, with lower connectivity between the PCC and MPFC and the right hippocampus (Figure 3D, Figure 4A). At 7T, the difference between the mean L-TLE and R-TLE Cohen’s *d* distributions was 0.51 (*p_FDR_*=0.008) in the PCC, and 0.30 (*p_FDR_*=0.042) in the MPFC. For the same subjects scanned at 3T the difference between the mean of the distribution of *Cohen’s d* values between L-TLE and R-TLE was 0.26 (*p_FDR_*=0.680) in the PCC and 0.35 (*p_FDR_=*0.160) in the MPFC. Thus, while the differences between distributions were in the same direction as at 7T, the hippocampo-DMN hypoconnectivity ipsilateral to the SOZ was higher at 7T than at 3T for the same subjects. In the extended 3T cohort, the Cohen’s *d* maps for L-TLE and R-TLE had a difference of 0.36 (*p_FDR_*=0.360) in the PCC, larger than that of the paired cohort at 3T, but not statistically significant, and still smaller than the difference in the paired cohort at 7T. The extended 7T cohort had a Cohen’s d map difference in the PCC of 0.42 (*p_FDR_*=0.048), larger than either of the 3T cohorts, but smaller than the original paired 7T cohort. We included the supplementary motor area (SMA) as a control region, and we did not see significant differences between L-TLE and R-TLE in cohort or field strength.

**Figure 3.**
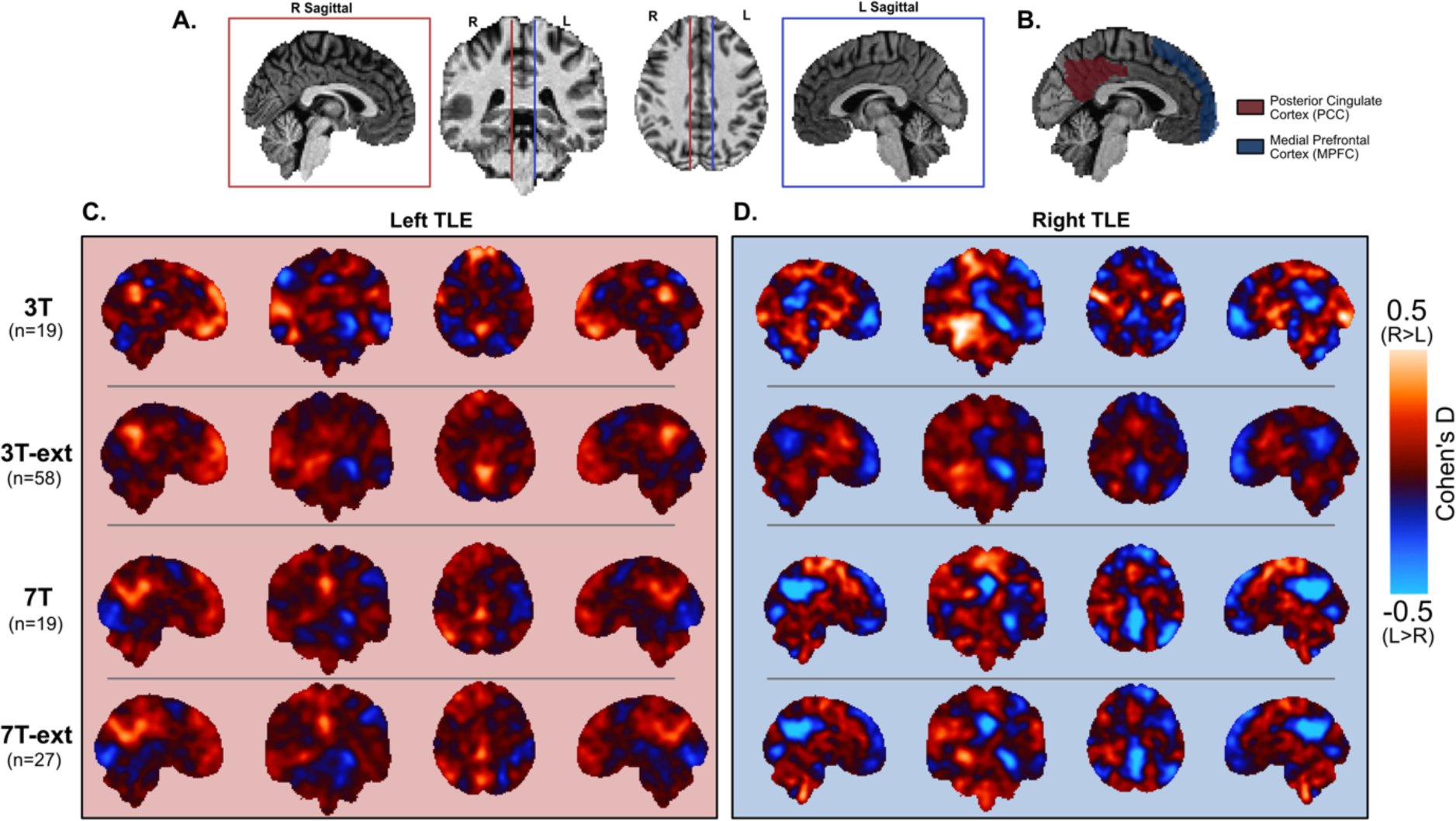
– Smoothed Voxelwise Cohen’s *d* Values. **Panel A** Shows the sagittal, coronal, and axial slices that are used in all the other panels of the figure. The red and blue lines represent the locations of the right and left sagittal slices respectively. **Panel B** shows the sagittal view of the posterior cingulate cortex (PCC) and the medial prefrontal cortex (MPFC) ROIs used in the study. **Panels C** and **D** show the smoothed (spatial Gaussian filter with σ = 2) voxel-wise Cohen’s *d* maps between the grouped left and right seed-to-voxel maps for left and right TLE respectively. The first, second, third and fourth rows of **C**. and **D.** show the maps corresponding to the paired 3T, the extended 3T, the paired 7T and the extended 7T cohorts respectively. 3T – paired 3T cohort, 7T – paired 7T cohort, 3T-ext. – extended 3T cohort, 7T-ext. – extended 7T cohort.

**Figure 4.**
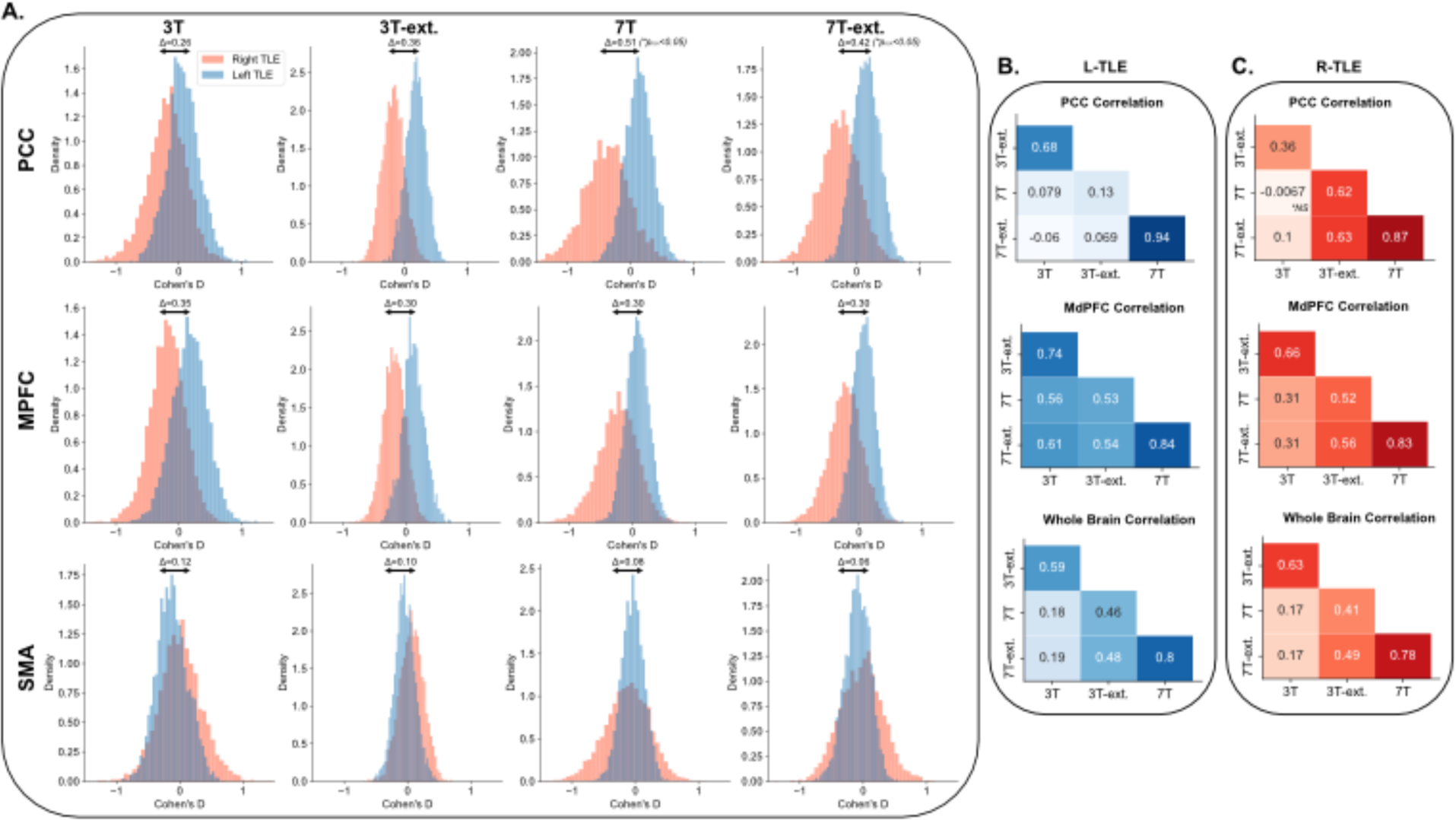
Voxelwise Cohen’s *d* Distribution in DMN Nodes. **Panel A**, top row, shows the distribution of voxelwise *Cohen’s d* values (from Figure 3) within the PCC ROI for (from left to right) the paired 3T, the extended 3T, the paired 7T and the extended 7T cohorts. Middle and bottom rows show the same distributions but for the MPFC and the supplementary motor area (SMA), the latter used as a control region. Statistically significant differences between distributions are specified in the figure. **Panels B** and **C** show the voxel-wise Pearson correlation between pairs of Cohen’s *d* maps for L-TLE (**Panel B**) and R-TLE (**Panel C**). Top, middle and bottom show the correlation within the PCC, MPFC and a whole brain mask. All correlations were statistically significant (*p_FDR_*<0.05) unless otherwise specified. NS: not statistically significant.

### Small sample sizes at 7T replicate findings in large samples at 3T

To determine if increasing the sample size at 3T makes findings more consistent to those seen at 7T, we computed voxel-wise correlations between Cohen’s d maps of the paired and extended 3T and 7T cohorts (Figure 4B-C). In R-TLE patients (Figure 4C), we saw in the PCC that the extended 3T cohort had a much better spatial correlation with the paired (Pearson’s r = 0.62, *p_FDR_*<0.001) and extended (Pearson’s r = 0.63, *p_FDR_*<0.001) 7T cohorts, compared to the original correlation between the paired 3T and 7T cohorts (Pearson’s r = -0.008, *p_FDR_*=0.10). That is, as we increased the number of subjects in the R-TLE group scanned at 3T, we saw findings more consistent to those seen at 7T. This pattern was also observed in the MPFC and whole brain, although to a lesser extent. For L-TLE (Figure 4B), correlations between the extended 3T cohort and the paired and the extended 7T cohorts remained low if the original paired 3T-7T correlation was low (as in the PCC), or high if the original paired 3T-7T correlation was high (as in the MPFC). For the whole brain correlation, we saw a pattern identical to that of R-TLE. The findings in the extended 3T cohort suggest that larger sample sizes at 3T lead to spatial distributions of effects that are more consistent with those seen at a much smaller sample size at 7T.

### Hippocampo-DMN connectivity at 7T lateralizes the SOZ in TLE better than at 3T

To quantify the ability of 3T and 7T rs-fMRI in lateralizing the SOZ we used the hippocampo-PCC connectivity at a single subject level by computing the mean value of the difference between smoothed right and left seed-to-voxel hippocampal maps within the posterior cingulate cortex (PCC) (Figure 5). Using 7T rs-fMRI (Figure 5A), the right-left hippocampus-PCC connectivity difference was significantly higher for L-TLE than R-TLE (p<0.001, Cohen’s *d* = 2.42). This demonstrates hypo-connectivity between the left hippocampus and the PCC in L-TLE, and hypo-connectivity between the right hippocampus and the right PCC in R-TLE, consistent with the group-level findings. At 3T, for the same subjects (Figure 5B), the right-left hippocampus-PCC connectivity difference was not significantly different between L-TLE and R-TLE (p=0.206, Cohen’s *d* = 0.62). Using the right-left hippocampus-PCC connectivity difference as a classification metric for separating L-TLE and R-TLE, we achieved superior lateralization of the SOZ at 7T (ROC AUC=0.97, 95% CI: 0.92-1.00) compared to 3T (ROC AUC=0.67, 95% CI: 0.36-0.98). These findings demonstrate superior SOZ lateralizing ability using the hippocampo-PCC (and hence hippocampo-DMN) at 7T than at 3T.

**Figure 5.**
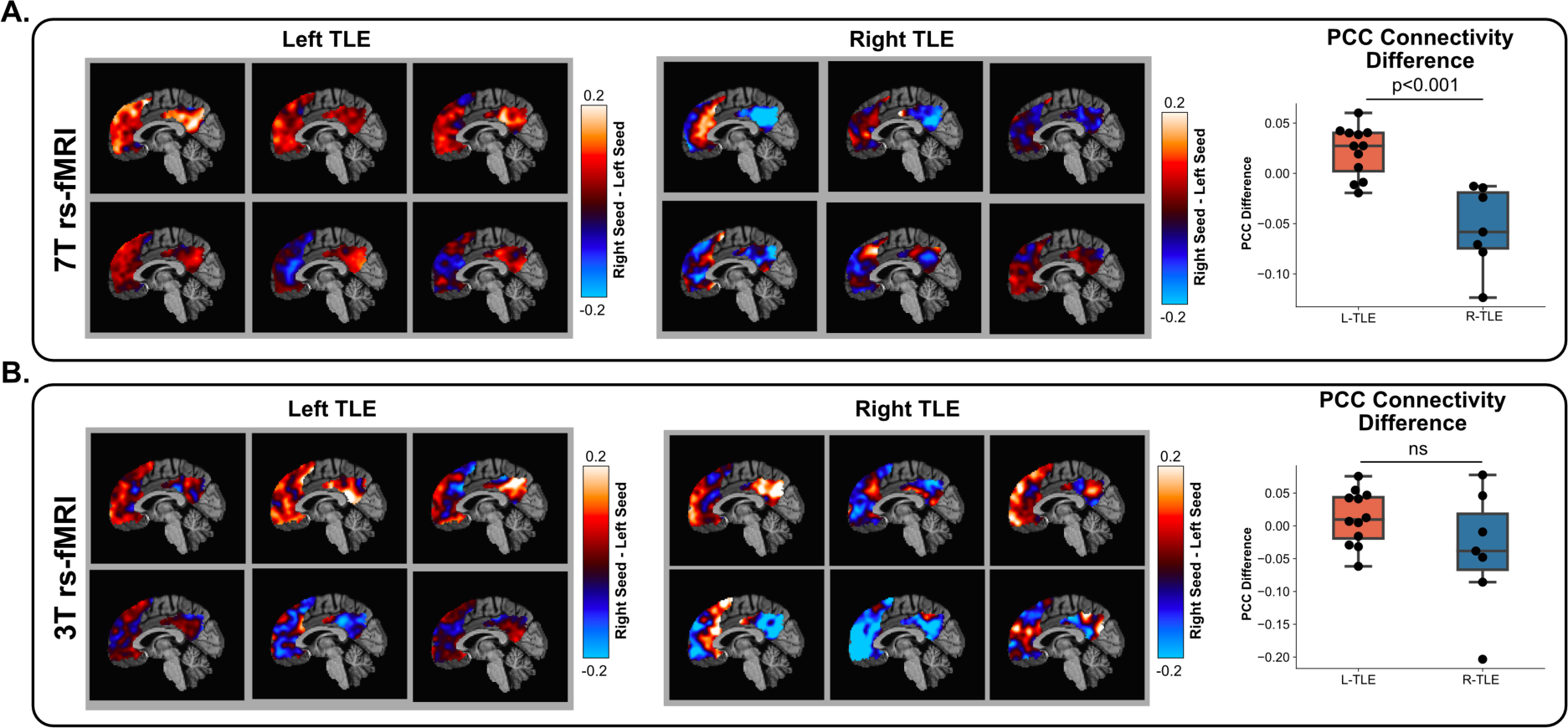
Comparing Left and Right Seed-to-Voxel Maps at a Subject Level. **Panels A** and **B** correspond to the paired 7T and 3T cohorts, respectively. On the left, the difference between right and left (right minus left) hippocampal seed-to-voxel maps for the same 6 left TLE subjects is shown at 7T (**Panel A**) and 3T (**Panel B**) field strengths. On the middle, the difference between right and left (right minus left) hippocampal seed-to-voxel maps for the same 6 right TLE subjects is shown at 7T (**Panel A**) and 3T (**Panel B**) field strengths. Only values within default mode network (DMN) ROIs are shown. On the right, boxplots represent the mean value of this difference inside the posterior cingulate cortex (PCC) ROI for all paired subjects. L-TLE: left TLE. R-TLE: right TLE. NS: not statistically significant.

### rs-fMRI Lateralization ability at 7T is independent of MRI lesional status

We tested whether the ability of 7T rs-fMRI to lateralize the SOZ in TLE was contingent on the presence of structural abnormalities on standard-of-care clinical MRI (MRI lesional status). We used the same right-left hippocampus-PCC connectivity difference from the previous section. At 7T, the separation between L-TLE and R-TLE (Figure 6A) remained regardless of whether there was an obvious structural abnormality (lesional, Cohen’s *d =* 2.94) or not (non-lesional, Cohen’s *d =* 1.83). At 3T (Figure 6B), an apparent separation between L-TLE and R-TLE was evident in the lesional sub-group (Cohen’s *d* = 2.32), but not in the non-lesional sub-group (Cohen’s *d* = 0.32). This preliminary evidence suggests that 3T rs-fMRI my be able to lateralize TLE in lesional cases but not in non-lesional cases, whereas 7T rs-fMRI’s lateralizing ability might be independent of lesional status.

**Figure 6.**
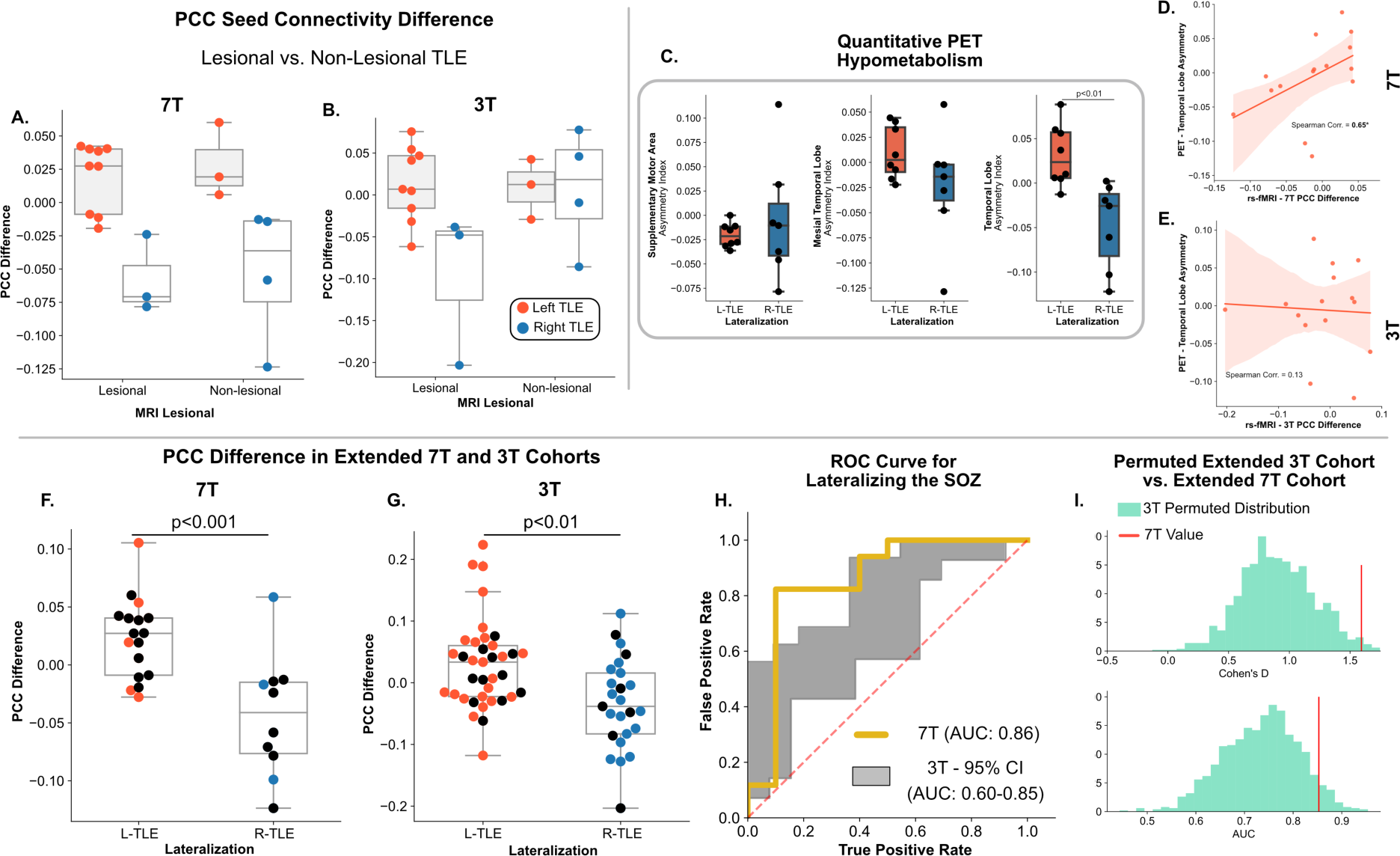
Effects of Lesional Status, PET Hypometabolism and Sample Size on Lateralizing Ability. **Panels A** and **B** show the mean difference between left hippocampal seed-to-voxel and right hippocampal seed-to-voxel maps within the PCC of subjects in the paired 7T and paired 3T cohorts respectively. Each panel has subjects subdivided based on SOZ laterality and the presence (lesional) or absence (non-lesional) of a structural abnormality on MRI. **Panel C** shows the quantitative FDG-PET contrast asymmetry (right-left/right+left) for (left) supplementary motor area, (middle) medial temporal lobe, and (right) temporal lobe across groups. **Panel D and E** show the correlation between the FDG-PET temporal lobe asymmetry and the right-left hippocampus-PCC connectivity difference metric measured at (D) 7T and (E) 3T. **Panels F** and **G** show the same as panels **A** and **B**, but for the extended 7T and 3T cohorts respectively. Black circles represent subjects from the paired 3T-7T cohort. **Panel H** shows the receiver-operating characteristics (ROC) curve for using the difference between left hippocampal seed-to-voxel and right hippocampal seed-to-voxel maps within the PCC as a metric for classifying L-TLE and R-TLE in the extended 3T and 7T cohorts. The gray area represents the 95% confidence interval ROC generated by resampling the extended 3T cohort to the sample size of the extended 7T cohort. Similarly **Panel I** (top) shows the Cohen’s d distribution between L-TLE and R-TLE in **Panels F** and **G**, (bottom) as well as the area under the curve (AUC) from **Panel H**, after resampling the extended 3T cohort to the same sample size as the extended 7T cohort. The value of the extended 7T cohort for each of these metrics is shown as a vertical red line.

### PET temporal hypometabolism correlates with findings at 7T

Since FDG-PET remains the neuroimaging clinical standard for SOZ lateralization in TLE^47^, we compared quantitative FDG-PET hypometabolism to our rs-fMRI findings at 3T and 7T. We measured the asymmetry index of the FDG-PET contrast within temporal and mesial-temporal ROIs. As expected, FDG-PET demonstrated temporal hypometabolism ipsilateral to the SOZ in both left TLE (positive asymmetry) and right TLE (negative asymmetry), causing a significant difference in asymmetry between groups (*p_FDR_* =0.009, Cohen’s *d =* 1.90) (Figure 6C). No significant FDG-PET asymmetries were detected in the supplementary motor area, nor in the mesial temporal lobe. We found a strong association between the FDG-PET hypometabolism, as quantified by the asymmetry in FDG-PET contrast, and the right-left hippocampus-PCC connectivity difference metric measured at 7T (Spearman Rho**=**0.65, p=0.01), but not at 3T (Spearman Rho**=**0.13, p=0.69) for the same subjects.

### Effect of sample size on subject lateralization

Since small sample sizes can both underestimate or overestimate measured effect sizes, we tested whether we could still lateralize the SOZ in the unpaired extended cohorts of TLE subjects scanned at 3T (n=62) and 7T (n=27). Findings in the extended cohorts replicated those in the original paired cohort, with the right-left hippocampus-PCC connectivity difference between L-TLE and R-TLE being significantly different at 7T (p<0.001, Cohen’s *d* = 1.59) (Figure 6E). At 3T, we also saw statistically significant differences between L-TLE and R-TLE (p=0.001, Cohen’s *d* = 0.88) (Figure 6F), although the effect size was much lower than at 7T. Since the extended 3T cohort had more subjects than the extended 7T cohort, we also generated 1000 random subsets of the extended 3T cohort with the sample size of the extended 7T cohort. The Cohen’s *d* in the extended 7T cohort (Cohen’s *d* = 1.59) was higher than in 96% of the permuted equal sized subsets of the extended 3T cohort (Figure 6I). The ability to lateralize the SOZ using the right-left hippocampus-PCC connectivity difference as a metric, as with our paired 3T-7T cohort, was higher at 7T (ROC AUC 0.85, 95% CI: 0.66-1.00) for the extended 7T cohort, compared to at 3T (ROC AUC 0.73, 95% CI: 0.59-0.86).

## Discussion

In this study we examined the effect of magnetic field strength (3T versus 7T) on measuring resting-state functional connectivity between the hippocampus and default mode network (DMN) nodes in patients with temporal lobe epilepsy (TLE). We found that while differences in connectivity patterns between L-TLE and R-TLE were in the same direction for 3T and 7T, the effect size was much larger at 7T for the same subset of subjects. However, when the sample size was increased in the 3T cohort, the findings became more consistent with those seen at 7T. We also demonstrated that the use of the hippocampal-posterior cingulate cortex (PCC) connectivity as a metric for lateralizing the seizure onset zone (SOZ) showed superior performance at 7T than at 3T. These results show that larger sample sizes at 3T may be able to replicate findings obtained at 7T, but 7T has better SOZ-lateralizing ability in TLE regardless of sample size. Finally, our findings also showed consistent results between the current neuroimaging standard FDG-PET and 7T, but not 3T rs-fMRI derived measurements.

The superior performance of 7T can be partially explained by the increased subcortical tSNR in 7T acquisitions when compared to 3T. The higher subcortical tSNR at 7T could have been a driver for higher quality (less noisy) hippocampal seed-to-voxel maps, allowing for stronger measured effects in the PCC and MPFC. Increased noise in the measurements at 3T could also explain why we did not see the same effects at 3T for the same subjects. Since noise decreases as the sample size increases, by increasing the sample size of the 3T cohort we were able to better replicate the findings at 7T. Thus, 3T fMRI might still capture similar effects as 7T rs-fMRI, but requires much larger sample sizes. Notably, tSNR remains an imperfect measurement of fMRI signal quality, particularly in clinical cohorts, where signal variance could be additionally influenced by pathological processes ^34, 35^. Furthermore, tSNR is highly dependent on scanner and acquisition parameters, making it difficult to compare across acquisition protocols.

Our results also corroborate prior findings regarding changes in hippocampal connectivity to the DMN in epilepsy, and in particular, TLE^15–20^. However, much of this past work has focused on group level analyses, making it challenging to assess the potential for clinical translation. We provide evidence that these DMN connectivity findings in TLE can replicated, and potentially expanded, at 7T, allowing for stronger effect sizes at a group level, and increased clinical utility at a single subject level. Despite the widespread availability of fMRI, and convergent evidence showing its potential in lateralizing epilepsy, the integration of fMRI into the presurgical pipeline remains limited. Some epilepsy centers use task-based fMRI to localize the eloquent cortex during presurgical planning^1^, but other clinical uses remain limited. We maintain that the lateralized changes we observed in the PCC for left and right TLE support the applicability of fMRI in pre-surgically lateralizing epilepsy. Furthermore, the superior performance of 7T in classifying left and right TLE could provide an avenue for fMRI to be more widely used during presurgical evaluation for seizure focus lateralization purposes, complementing the findings of other functional imaging modalities such as MEG, PET and SPECT.

The main limitation of our study is the sample size of our paired 3T-7T cohort. Increasing the sample size of the paired cohort would allow to better understand the differences between 3T and 7T fMRI acquisitions in epilepsy and may allow a better exploration of regions/networks of interest other than the DMN. We also believe that larger sample paired 3T-7T cohorts would allow us to test whether there is an upper limit on how much 3T can approximate the findings of 7T by simply increasing the sample size. In other words, one possibility is that 7T can find effects that cannot be observed at 3T regardless of how large the sample size of the 3T cohort is. Future studies should focus on acquiring a larger paired 3T-7T dataset to better answer these questions. Relatedly, another limitation of our study is that we only focused on changes within a defined brain region, the DMN, and specifically the PCC and MPFC. However, some of our findings (the superior subcortical tSNR at 7T, particularly in the thalamus), might suggest that there is an avenue to further explore the subcortical connectivity profile of epilepsy using ultra-high field imaging. Our group has started to explore this in past studies^21^, but these were limited to the hippocampus. With larger samples in a 7T only cohort, we plan on further exploring mesial temporal and subcortical functional relationships in TLE and other epilepsy subtypes.

## Conclusion

In this study we compared rs-fMRI derived measurements from 7T and 3T in TLE patients. We demonstrated that for the same subjects, studying hippocampal connectivity to the DMN yielded larger effect sizes with 7T rs-fMRI than with 3T rs-fMRI. We also demonstrated that by leveraging this increased effect size, 7T rs-fMRI can outperform 3T rs-fMRI in lateralizing the SOZ in TLE. This study provides support for the use of 7T rs-fMRI in epilepsy presurgical evaluation, and shows that 7T rs-fMRI can help better characterize functional network alterations in TLE at smaller sample sizes than those required for 3T fMRI.

## Data Availability Statement

The processed paired 3T and 7T neuroimaging data and methodological pipeline used in this study will be made available on a public data repository (OSF) upon publication of the manuscript. The repository will be made publicly accessible to ensure that the data and methods used in this study can be transparently and independently verified.

## Acknowledgements

AL and KAD received support from NINDS (R01NS116504). NS received support from American Epilepsy Society (953257) and NINDS (R01NS116504).

## Author Contributions

**Alfredo Lucas**: Conceptualization, Methodology, Software, Validation, Formal analysis, Writing - Original draft preparation. **Eli J. Cornblath**: Conceptualization, Writing - Review & Editing, Data curation. **Nishant Sinha**: Conceptualization, Writing - Review & Editing. **Lorenzo Caciagli**: Conceptualization, Writing - Review & Editing. **Peter Hadar**: Data Curation, Writing - Review & Editing. **Ashley Tranquile**: Data Curation, Writing - Review & Editing. **Joel M. Stein**: Data Curation, Conceptualization, Writing - Review & Editing. **Sandhitsu Das**: Data Curation, Conceptualization, Writing - Review & Editing. **Kathryn A. Davis**: Supervision, Project administration, Funding acquisition, Data Curation, Conceptualization, Writing - Review & Editing.

